# Colorectal Cancer Risk and Ancestry in Colombian admixed Populations

**DOI:** 10.1101/2023.03.02.23286692

**Authors:** Angel Criollo-Rayo, Mabel Elena Bohórquez, Paul Lott, Angel Carracedo, Ian Tomlinson, Jorge Mario Castro, Gilbert Mateus, Daniel Molina, Catalina Rubio Vargas, Carlos Puentes, CHIBCHA Consortium, Magdalena Echeverry, Luis Carvajal

## Abstract

Several colorectal cancer susceptibility disease loci have been discovered through Genome-wide association studies. However most of the variants were originally identified in Caucasian populations. Demographic history and admixture background may impact the association of known CRC variants due to the differences in linkage disequilibrium across different populations. We performed a genomic study in a sample of 955 cases and 968 controls from admixed populations in Colombia by genotyping ∼1 million SNPs aimed to detect the impact of genetic structure in the association of 20 known SNPs risk for colorectal cancer. The replication was reached for eleven out of 20 nominally associated SNPs; with allelic odds ratios (OR) between 1.14 and 1.41, indicating a minimal individual risk increment; on the other hand, the overall OR for co-inherited SNPs was 5.4 (95% CI: 3.052-9.731, *P*=1.16E-08). Most of the variants followed a recessive model with significant homozygous ORs distributed between 1.3 and 1.65. The major associated markers were: rs4939827 (18q21.1, *P* =7.35E-6), rs10411210 (19q13.11, *P*=0.001) rs10795668 (10p14, *P*=0.0024), rs4444235 (14q.2.2, *P*=0.005), rs961253 (20p12.3, *P*=0.006), rs16892766 (8q23.3, *P*=0.011) and rs1050547 (8q24.21, *P*=0.017). Additionally, European ancestral component was associated with colorectal cancer risk (*p=6*.*48E-04, OR = 4*.*244 95% IC: 1*.*701-10*.*68*). Our findings in Colombia indicates a significant contribution of the known CRC risk SNPs to the disease in the Colombian population, which in turns can be explained by the genetic European component influx during the admixture process. The unassociated SNPs indicates frequency and genetic structure differences between European and Colombian populations or due to the sample process.

## INTRODUCTION

Colorectal carcinoma (CRC) affects 1.85 million people around the world each year and represents almost 10.2% of the global cancer incidence burden (Mattiuzzi et al., 2019). Among the malignancies, CRC has exhibited one of the greatest increases in Colombia between the years 1984 and 2008, showing a trend in equalizing incidence and mortality rates (Bray & Piñeros, 2016) (Piñeros et al., 2013). An updated population-based cancer registry data in Colombia showed an age-standardized incidence rates between 8.4 and 16.2 (de Vries et al., 2020).

Colorectal cancer is a complex disease involving a dynamic interaction between environmental and genetic factors such as dietary/lifestyle and high/moderate-penetrance variants (Chapelle, 2004; Cross et al., 2010). Contributions from twin cohort analysis have estimated a large effect of heritability (35%) on CRC (Lichtenstein et al., 2000); from this fraction at least ∼5% account for familial hereditary syndromes caused by high impact mutations (Jasperson et al., 2010; Tomlinson, 2015), while the rest ∼30% of CRC cases that seems to be familial, could be mediated by a combination of common low penetrance variants (Short et al., 2015; Whiffin et al., 2013). This approach provides support for a polygenic model of disease susceptibility, with the co-inheritance of multiple genetic variants, each with a modest individual effect for CRC (M. G. Dunlop et al., 2012; Gafni et al., 2021; Thomas et al., 2020; Whiffin & Houlston, 2014). Genome-wide association studies have so far discovered several common susceptibility loci for CRC that contain tagging SNPs at 1q41, 3q26.2, 6p21.2, 8q23.3, 8q24.21, 10p14, 10q24.2, 11q13.4, 11q23.1, 12q13.12, 14q22.2, 15q13.3, 16q22.1, 18q21.1, 19q13.11, 20p12.3 and 20q13.33 (Malcolm G. Dunlop et al., 2012; Houlston et al., 2010; Tanskanen et al., 2018; Tenesa et al., 2008; Tomlinson et al., 2008; Tomlinson et al., 2007; Whiffin et al., 2014; Zanke et al., 2007), among others from Asian studies (Lu et al., 2019; Tanikawa et al., 2018). Most of these studies have been conducted in Caucasian-derived ancestry populations, and few follow-up replication studies have been addressed in Hispanic or Latin American populations. In consequence the effect of the known variants remains almost uncharacterized, with limited information about CRC genetic in Latino populations. Since there are variations in allele-frequencies and Linkage disequilibrium (LD) patterns across different continental populations, the impact of these variants needs to be investigated to ascertain their roll in CRC in Hispanic/admixed populations. For example, Colombian people from Andean region have originated mostly from the admixture process between Native Americans and Europeans during Colonial period, and it is well known that the extension of LD will depend on the number of generations since the admixture event and the particular recombination histories; the less generations the higher LD extension (Jobling et al., 2014; Winkler et al., 2010), which is also useful to detect chromosomal regions highly associated with disease phenotype by admixture mapping (Smith & O’Brien, 2005). On the other hand, it is important the characterization of putative genotype-phenotype correlations in Latino populations, due to the possible benefit in improving the efficacy of CRC management and prevention strategies for CRC by targeting people in greater risk. Thus, to further our knowledge and explore the published CRC risk regions, we addressed here the impact of genetic structure on the association of 20 known low penetrance loci in Colombian cases and controls with proven genome-wide significance considering ancestry among other variables.

## METHODS

### Study populations and sampling

The cases and controls were collected within the project *Genetic Study of Common Bowel Cancer in Hispania and the Americas* –CHIBCHA-(https://cordis.europa.eu/project/id/223678/reporting/es), in different health centers or cancer research institutions across the Colombian Andean region: Hospital Federico Lleras Acosta (Ibagué), National Cancer Institute (Bogotá D.C.), Pablo Tobón Uribe and San Vicente de Paul Hospitals (Medellín), Fernando Moncaleano Perdomo University Hospital (Neiva) and Population Registry of Cancer of Pasto (Pasto); among others as have been previously described (Bohórquez et al., 2016). A total of 1923 participants were included for the present study, distributed in 955 cases and 968 controls. The patients were prevalent cases under 75 years old diagnosed with colon adenocarcinoma/adenoma. Controls were unrelated individuals above 55 years old without any cancer diagnosis or personal family history of colorectal neoplasia (up to second-degree relatives). Recruited individuals received complete information about the study and signed an informed consent, information letter and filled out a socioeconomic interview. We Collect blood samples from cases and controls, as well as the clinical and pathological information of cases. This study was approved by the committee of Bioethics of the University of Tolima and as well as the collaborating institutions. The research protocol adhered to the principles of Helsinki declaration. The present study included individuals from 1000 genomes project: Africans (YRI=108) and Europeans (CEU=99 GBR=104, IBS=107); and also a Native American dataset from Esteban Parra (Tlapanec=5, Quechua-Peruanos=24, Mexicanos-Nahua/Mixtec/Maya=34, Aymara-Bolivia=25).

### DNA extraction, genotyping and QC

DNA was extracted from the cases and control samples using the automated equipment MAXWELL-16 (Promega) and quantified using spectrophotometric methods in a Nanodrop ND-2000. Two different microarrays were genotyped (Custom-550434 and LAT) for a total of 1,169,207 markers SNPs using the GenTitan Axyom Affymetrix platform at the University Santiago de Compostela. A DQC (Dish Quality Control, Affymetrix Axiom Array QC) was perform to the raw genotype data and dishes with < 0.82 values were removed from the analysis and samples with a genotyping rate <97% were subsequently excluded. Individual QC steps was performed in PLINK according to the following parameters: 1) sex check based on X-chromosome homozygosity rates (homozygosity in females > 0.2 and in men <0.8), 2) individual rate for missing genotypic data (Threshold = 0.021528474), 3) excess of heterozygosity thresholds (>0.2684462 and <0.2202541), 4) exclusion of genetically related individuals based on the identity paired matrix: IBS. Marker QC included steps aimed to exclude those that don’t meet the next criteria: 1) high rate of missing genotypic data (<95%), 2) out of Hardy-Weimberg equilibrium (P<0.001) and 3) low minor allele frequency (< 5%). A total of 1,169,944 SNPs passed QC filters for the analysis.

### Population structure and global ancestry

A set of 87.359 markers (set 87K) was filtered out in cases and controls by using PLINK (Purcell et al., 2007), we excluded regions in high LD such as HLA (Anderson et al., 2010; Price et al., 2008) or markers correlated with each other (r^2^ ≥ 0.2). This set of SNPs was obtained in order to filtered out common markers between Colombian, African, European and Native American datasets in PLINK. We implemented 87K set markers for the population genetic structure analysis calculating principal components (SMARTPCA) (Patterson et al., 2006), global ancestry proportions (ADMIXTURE) and Fst values (PLINK, HierfStat R package). In order to calculate the admixture membership proportions, we run ADMIXTURE starting with a k=2 up to k=5, with 100 replicates, including cases, controls and reference populations from the most probably ancestral origins: Native Americans, Europeans and Africans. The results were analyzed in R using non-parametric statistics (U-Mann Whitney) to compare cases v.s controls or Colombian subpopulations. In regards to PCA, eigenvalues and eigenvectors were plotted for each individual according to the calculated coordinates and the percentages of genetic variation explained by each component and analyzed using the Tracy-Widom test. The graphical analysis and plots for PCs, admixture proportions and histograms were performed in R language (Wickham, 2016). In order to quantify the magnitude of genetic differentiation between cases and controls or among Colombian populations, the Fst was calculated following the Wright method (Wright, 1949) and Weir-Cockerham (Weir & Cockerham, 1984), estimating a parameter for each autosomal diploid locus. By Cockerham’s (1984) definition, the Fst is the ratio of the variance between subpopulations, relative to the variance in the overall population.

### Association analysis

For the association analysis, we include 20 low and variable CRC risk susceptibility SNPs located in the following regions: chromosomes: 1q41 (rs6691170, rs6687758), 3q26.2 (rs10936599), 6p21.2 (rs1321311), 8q23.3 (rs16892766), 8q24.21 (rs10505477, rs7014346, rs6983267), 10p14 (rs10795668), 10q24.2 (rs1035209), 11q13.4 (rs3824999), 11q23.1 (rs3802842), 12q13.12 (rs11169552), 14q22.2 (rs4444235), 15q13.3 (rs4779584), 16q22.1 (rs9929218), 18q21.1 (rs4939827), 19q13.11 (rs10411210), 20p12.3 (rs961253) and 20q13.33 (rs4925386). Each one of these markers are tagging specific chromosomal regions previously published (Malcolm G. Dunlop et al., 2012; Houlston et al., 2010; Albert Tenesa et al., 2008; Tomlinson et al., 2008; Tomlinson et al., 2007; Whiffin et al., 2014; Zanke et al., 2007). The association study was carried out using PLINK and R language (Version 3.2.1; http://www.rproject.org/) (R Core Team, 2021). Initially, the complete association models were evaluated: allelic, genotypic, dominant and recessive; and statistical significance was assessed by implementing two-tailed P values and corrections for multiple trials (Bonferroni, False Discovery Rate: FDR, among others). Two degrees of freedom were used for the genotypic models and one for the others (chi squared test). Additionally, logistic regression models were implemented for CRC susceptibility based on the allele risk dose for each SNP; for which covariates such as the first four principal components (PC1, PC2, PC3 and PC4), socioeconomic stratum, level of education and global ancestry were considered. For multivariate models in which Native American and European ancestral proportions were included, the variance inflation factor (VIF) was calculated due to the correlation between ancestry and principal components (John & Sanford, 2011). Subsequently, one of the covariates with a VIF > 5.0 was excluded, according to the desired model. On the other hand, a cumulative risk analysis was developed, generating categories based on the number of risk alleles carried and the percentage of individuals in each one. The cumulative effect on the susceptibility to CRC was measured through logistic regression models taking the lowest risk category as the reference group or comparing cases and controls in each one.

## RESULTS

The CHIBCHA consortium recruited a total of 1923 individuals (955 cases and 968 controls) in Colombia that were genotyped for 1’036.937 SNPs. The sampling process was focused across the Andean Region, where most of patients (98.5%) and controls (99.4%) were born according to reportedly interview. The clinical and histological characteristics for the study cohort have been previously well described (Bohórquez et al., 2016). In TABLE 1 we included socio-demographic information such as social class, education level and gender as covariates for statistical models. According to the TABLE 1, at least a ∼83% of cases and ∼77% controls had a low (primary) or middle (secondary/technical) education level. Most of cases and controls (77 and 76% respectively) were distributed in the low/medium Social class levels.

**TABLE 1:**
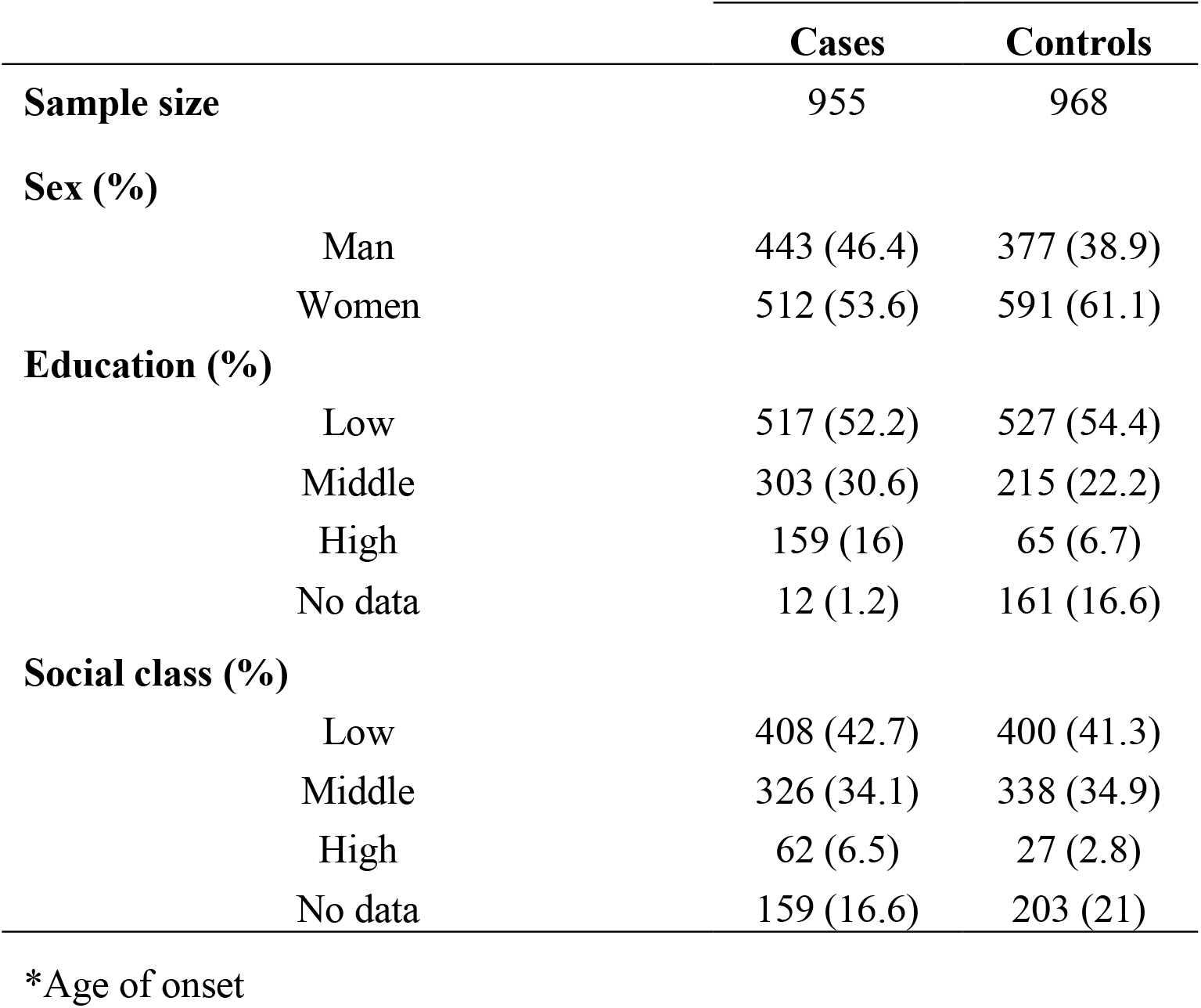
Socio-economic, demographic and characteristics of the colorectal cancer cases and controls in CHIBCHA-Colombia.

### Genetic population structure and CRC risk

The population structure was obtained by two different methods applied in ADMIXTURE and SMARTPCA. PCA figure show a high individual variation in cases and controls from the study (FIGURE 1), depicting the same pattern of population structure, mostly distributed between Native Americans and Europeans (FIGURE 1a), which agrees with the main ancestral background in the Andean region, while PC1 splits African and non-African populations.

**FIGURE 1.**
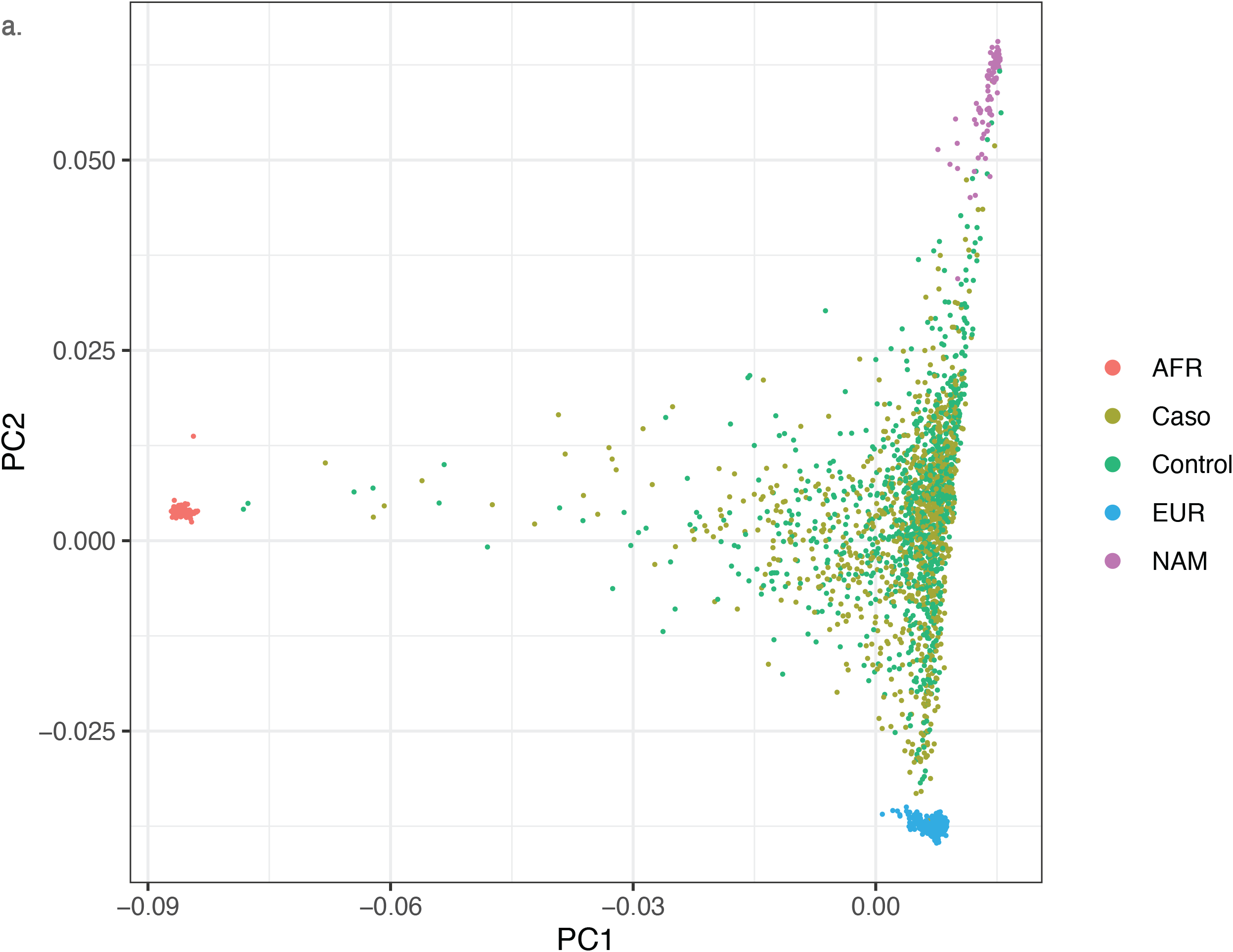
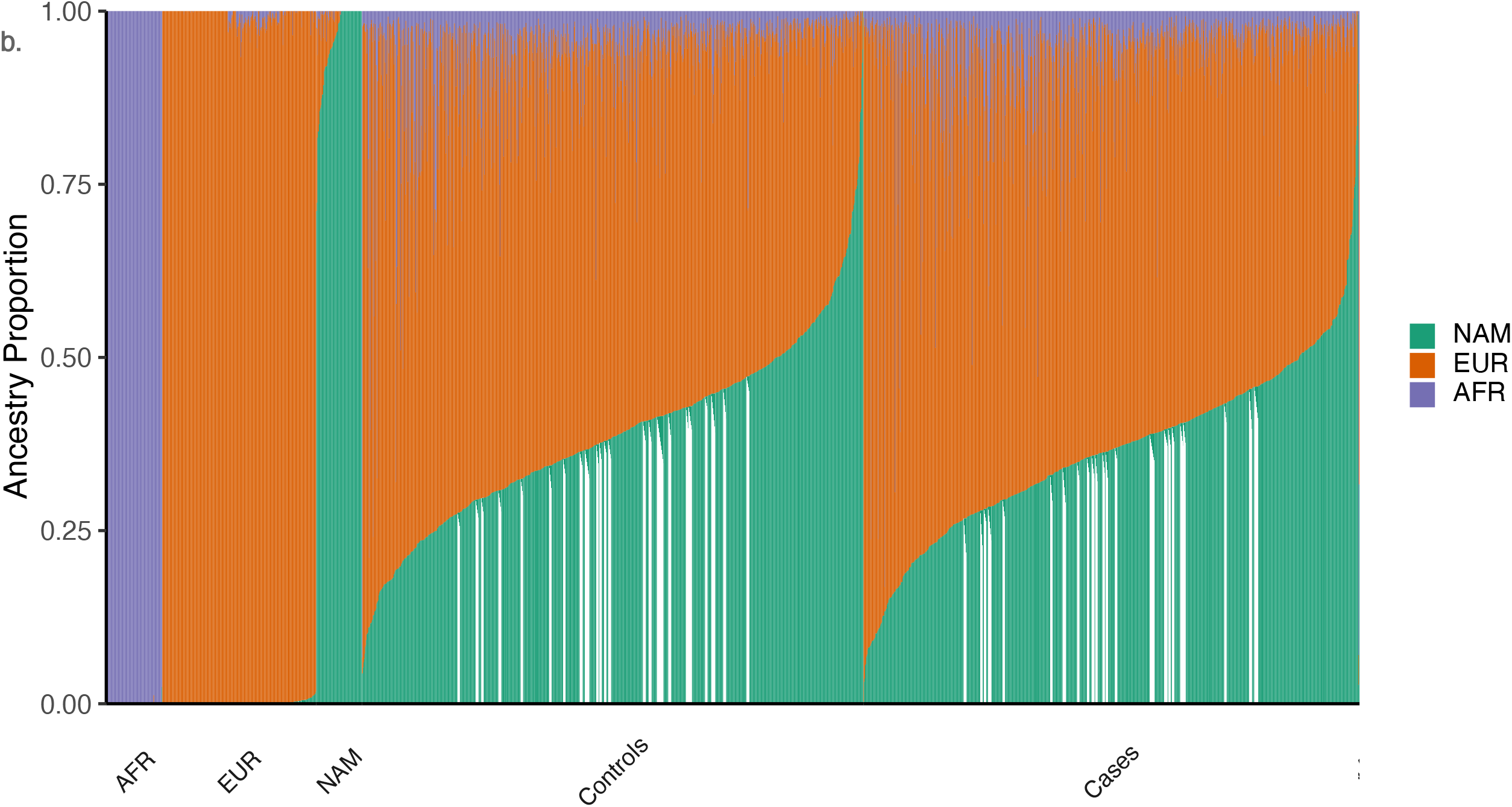
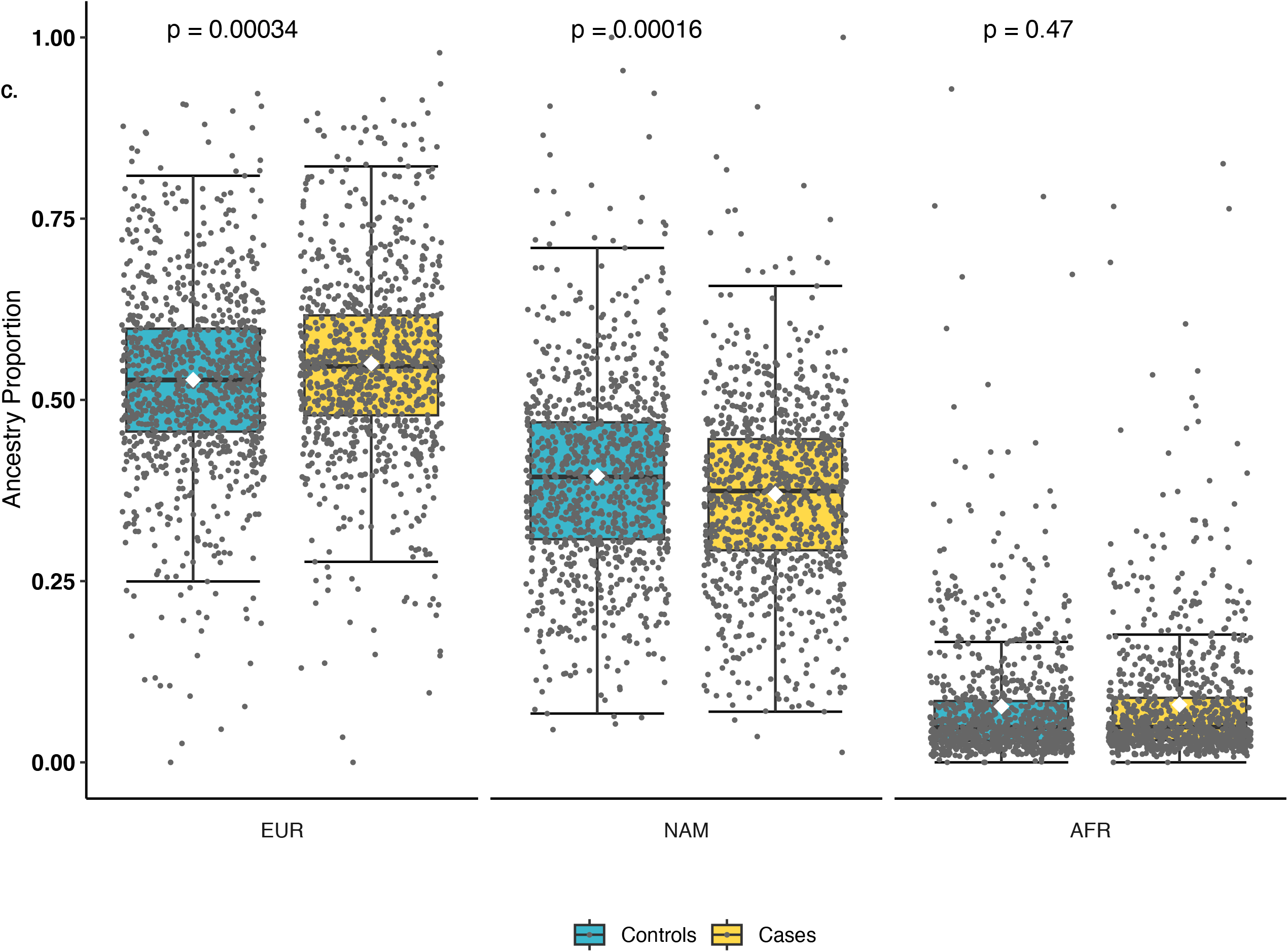
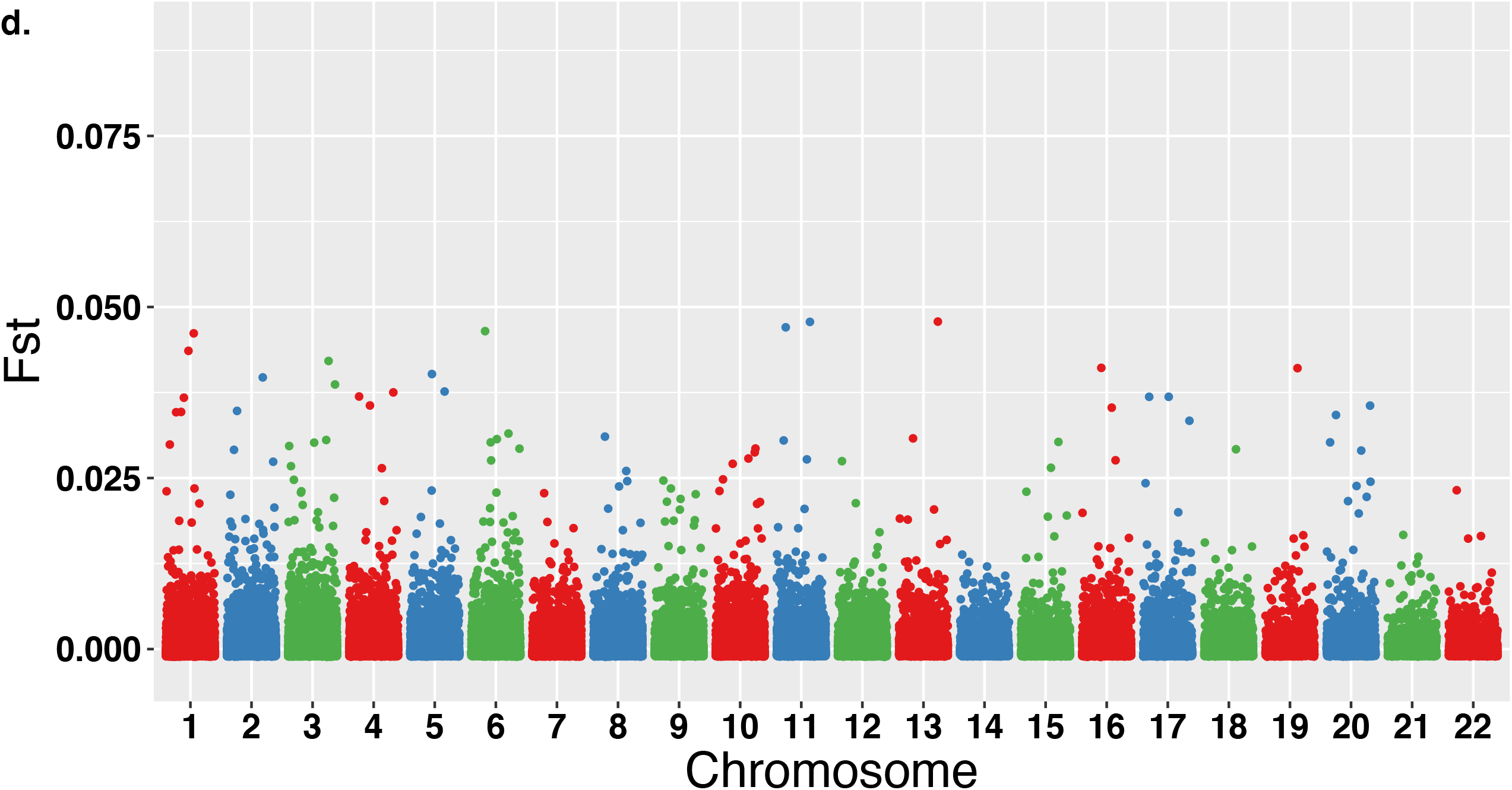
Different approaches to detect genetic population structure in the sample of cases and controls. a. Principal components analysis of in the in 955 cases of CRC and 968 controls of the Colombian sample using 87K SNPs, including the reference populations from African, European and Native American origins. b) Admixture was ascertained by using ADMIXTURE V2 with supervised models, including the reference European (EUR) or African (AFR) samples from 1000 genomes project and Native American (NAM) data from Esteban Parra. Admixture proportions memberships were analyzed comparing a) cases and controls. c) box-plot for the three admixture proportions considered for the present work (NAM, EUR and AFR). d) Global genomic distribution of *Fst* by marker, in the set of 87k SNPs in the Colombian sample along the 22 autosomal chromosomes. a) Fst values for each SNP comparing cases and controls (Weighted *Fst=* 0,000234, mean Fst=0.000359).

In order to explore the variable pattern of the observed genetic diversity, we performed an admixture analysis (FIGURE 1-b,c). In regards to ancestral membership proportions, controls had higher Native American ancestry (x^−^= 0.39±0.13), compared with CRC cases (x^−^= 0.37±0.126); using a non-parametric test (Mann-Whitney U) significant differences were observed (p = 0.0001621). The European ancestral component behaved in the opposite way, being significantly higher in the cases (x^−^= 0.550±0.13) than in controls (x^−^= 0.527±0.13) (p = 0.0003423). African ancestry in both, cases and controls was ∼8% (s.d = 0.09). Most regions sampled in Colombia showed similar pattern (TABLE 2).

**TABLE 2:** Admixture proportions for colorectal cancer cases and controls along different sampled regions in Colombia.

| Ancestry | Region | Controls | Cases | P |
| --- | --- | --- | --- | --- |
| **EUR** | Nariño | 0.386 ± 0.140 | 0.460 ± 0.121 | 0.003411 |
|  | Boyaca | 0.479 ± 0.092 | 0.510 ± 0.075 | 0.1003 |
|  | Huila | 0.508 ± 0.101 | 0.519 ± 0.109 | 0.3582 |
|  | Cundinamarca | 0.524 ± 0.096 | 0.533 ± 0.094 | 0.4006 |
|  | Tolima | 0.515 ± 0.104 | 0.519 ± 0.096 | 0.7717 |
|  | Santanderes | 0.541 ± 0.073 | 0.570 ± 0.104 | 0.01245 |
|  | Valle_y_Cauca | 0.441 ± 0.155 | 0.454 ± 0.179 | 0.09194 |
|  | Eje_cafetero | 0.580 ± 0.109 | 0.613 ± 0.120 | 0.6101 |
|  | Costa | 0.516 ± 0.146 | 0.487 ± 0.138 | 0.01613 |
|  | Antioquia | 0.587 ± 0.141 | 0.620 ± 0.139 | 0.001728 |
| **NAM** | Nariño | 0.567 ± 0.158 | 0.497 ± 0.129 | 0.003411 |
|  | Boyacá | 0.496 ± 0.096 | 0.461 ± 0.079 | 0.1003 |
|  | Huila | 0.440 ± 0.105 | 0.420 ± 0.102 | 0.3582 |
|  | Cundinamarca | 0.434 ± 0.103 | 0.420 ± 0.094 | 0.4006 |
|  | Tolima | 0.421 ± 0.104 | 0.423 ± 0.096 | 0.7717 |
|  | Santander | 0.421 ± 0.069 | 0.382 ± 0.107 | 0.01245 |
|  | Valle | 0.399 ± 0.150 | 0.355 ± 0.145 | 0.09194 |
|  | Eje Cafetero | 0.343 ± 0.099 | 0.322 ± 0.097 | 0.6101 |
|  | Costa | 0.250 ± 0.099 | 0.336 ± 0.111 | 0.01613 |
|  | Antioquia | 0.301 ± 0.109 | 0.271 ± 0.097 | 0.001728 |
| **AFR** | Nariño | 0.047 ± 0.113 | 0.042 ± 0.040 | 0.003411 |
|  | Boyacá | 0.025 ± 0.014 | 0.030 ± 0.020 | 0.1003 |
|  | Huila | 0.052 ± 0.023 | 0.062 ± 0.035 | 0.3582 |
|  | Cundinamarca | 0.042 ± 0.047 | 0.047 ± 0.049 | 0.4006 |
|  | Tolima | 0.064 ± 0.061 | 0.058 ± 0.036 | 0.7717 |
|  | Santanderes | 0.037 ± 0.029 | 0.047 ± 0.052 | 0.01245 |
|  | Valle y Cauca | 0.160 ± 0.211 | 0.191 ± 0.220 | 0.09194 |
|  | Eje Cafetero | 0.076 ± 0.051 | 0.065 ± 0.054 | 0.6101 |
|  | Costa | 0.234 ± 0.114 | 0.177 ± 0.126 | 0.01613 |
|  | Antioquia | 0.112 ± 0.087 | 0.109 ± 0.086 | 0.001728 |

The entire sample of cases and controls showed mostly Native American and European ancestry admixture components, both of them with the same distribution pattern close to normal distribution (logistic), while African ancestry showed a gamma pattern (symmetry=3,72 and curtosis= 23,2), this means a low variation around the mean and bias to the higher African proportions (supplementary figure 1). The probability density distribution curve indicates that some regions from Colombian Andes, have a higher African ancestral component which decay soon to lower levels, indicating a recent admixture due to human internal diasporas, mostly from Afrocolombians from Pacific or Atlantic coast to the central regions. Complementary analysis based on Fst statistics showed a low differentiation between cases and controls with the 87K SNPs dataset (FIGURE 1-d) (Weighted *Fst=* 0,000234, mean Fst=0.000359) or 246K (Weighted *Fst=* 0,000253, mean *Fst*=0.000357). These results are important when considering population substructuring for association analyses, since large differences in allele frequencies between cases and controls could generate false associations.

### Relationship between the global ancestral component and the risk of CRC

Given the significant differences observed between colorectal cancer cases and controls form the study (FIGURE 1-c), different logistic regression models were performed in order to test the association between Native American or European ancestry with CRC susceptibility. A first approach consisted in generating a univariate basal model (*status* = *β*_0_ + *β*_1_ * *ancestry + ε*_*i*_), taking as the dependent variable the case/control status and as the independent or predictor variable one of the ancestries at a time, due to the multicollinearity between native, European and African ancestries (*R*^*2*^ = -0.764, *95% IC: -0*.*782 a -0*.*744, p< 2*.*2e*^*-16*^).). The autocorrelation was measured through the variance inflation factor (VIF), in order to avoid the overestimation of the regression coefficients. The basal model showed a significant association for European (*estimated= 1*.*45 ± 0*.*399, p = 0,000267; OR=4*.*274, IC 95%: 1*.*97-9*.*39*) and Native American ancestry (*estimated = -1*.*505 ± 0*.*39, p = 0*.*000114; OR = 0*.*222, IC 95%: 0*.*103 - 0*.*475*). When both ancestries were included in a single model, there was an standard deviation increment for the stimates as well as VIF=2.33 (1<VIF<5, moderate autocorrelation). Additional models included covariates such as social class, education level, sex and geographical origin of the sample in order to correct for bias. We evaluated a total of five different models (Suplementary Table 1), which resulted significant for *Wald* test (H_0:_ *β* = 0) for the European ancestry coeficient stimates (*m1= 0*.*00027, m2=2e-04, m3=2*.*5e-05, m4=0*.*00016, m5=0*.*002*). Model goodness of fit was ascertained by maximum likelyhood ratio testing between pairs of models, in this way model 5 (m5, *status* = *β*_0_ + *β*_1_ * *EUR* + *β*_2_ * *Education* + *β*_3_ * *origin* + *β*_4_ * *social* – *class* + *β*_5_ * *sex* + *ε*_*i*_) had the best goodnes of fit (*p=6*.*48E-04, LogLik= -1003*.*1*) and *OR = 4*.*244* (*95% IC: 1*.*701-10*.*68*). Native American ancestry was found to be negatively associated with CRC risk when included the covariates already mentioned (*OR = 0*.*113, IC 95%: 0*.*039 - 0*.*316*).

If the increment of the European ancestral component is directly associated with CRC susceptibility, it is possible that some ranges are more associated than others; in this sense the variable was stratified by quartiles (ranges with 25% increments). The logistic regression was performed adjusting for the covariates already mentioned above, and taking as the baseline level to compare the lowest European ancestry range between 0 and 25% (dummy contrast). In the first strategy the basal level was compared with the highest ones, the most significant ranges were 5-75% (P= 0.049 y) and 75-100% (P= 0.0407), presenting adjusted ORs of 2.30 (IC95%: 1.019-5.44) and of 2.64 (IC95%: 1.057-6.863) respectively. When Native American ancestry was analyzed in the same way, the significant range was between 50-75% (P=0.022) with an adjusted OR of 0.591 (IC95%: 0.376-0.926). This reflects the opposite effect, namely protective against the increase in risk that was observed for European. Some of the covariates turned out to be significant, such as, for example, socioeconomic stratum and educational level, as when ancestry was analyzed as a continuous or categorical variable.

### SNPs associated with CRC risk

After excluding SNPs that did not fit call-rate or HWE thresholds, we kept three sets of markers for the CRC known risk markers downstream analysis: (1) 20 known tagSNPs with 99.7 call-rate, (2) 3.788 genotyped and (3) 30.996 imputed SNPs for all the risk regions.

Previously we observed the association between the European genetic component and the risk for CRC, but a protective association was found for the Native American ancestry. However, the analyzed ancestry corresponds to global genomic behavior, which may have origins in the parental populations we considered; it was necessary to make a locus-specific variation study approach.

Two methodological approaches were used to assess the role of ancestry and locus-specific risk for CRC. An initial focus adressed the heritable genetic variation that increased susceptibility to CRC, specifically those markers previously reported in populations of Caucasian origin; although, the chromosomal regions where these variants lie in cases, can have either an excess (or not) of Native American, European or African ancestry irrespectively of the global ancestry risk. Therefore, we analyzed the haplotypic structure in these regions. A second approach consisted in the search for association of chromosomal regions with locus-specific ancestry significantly associated to CRC risk. So, we hypothesized the existence of specific chromosomal regions, with a high proportion of European or even Native American ancestry, well differentiated between cases and controls by using local ancestry and admixture mapping

### Association of 20 SNPs with CRC risk in Colombia

The association results for each of the 20 markers are reported in TABLE 3. Eleven out of 20 markers were significantly associated with CRC risk in the Colombian population (P < 0.05, without adjusting for multiple tests), with a magnitude effect in the same direction as in other populations. After the false discovery rate test (FDR), seven markers remained significant rs4939827 (18q21.1), rs10411210 (19q13.11), rs10795668 (10p14), rs4444235 (14q22.2), rs961253 (20p12.3), rs16892766 .3) and rs10505477 (8q24.21). Finally, Bonferroni correction The logarithmic-additive effect of each risk allele per SNP on CRC susceptibility (coded as 0, 1 or 2 alleles) was estimated. Each SNP was analyzed in a logistic regression model separately, adjusting for gender, principal components, educational level, and social stratum. Five of the 20 markers remained significant: rs4939827 (OR=1.25, 95% CI=1.067-1.456, P=0.00544), rs10411210 (OR=1.24, 95% CI=1.009-1.52, P = 0.0412), rs10795668 (OR= 1.19, 95% CI=1.015-1.393 P= 0.03216), rs4444235 (OR=1.20, 95% CI= 1.038-1.405, P= 0.01451) and rs16892766 (OR=1.52, 95% CI=1.082-2.142.5 P= 0.01), these five SNPs had already been found to be significant in previous tests. For these markers, the increase in the number of risk alleles is associated with CRC susceptibility, which is supported by the significant ORs in the association tests under recessive models, in which the homozygous status of the risk allele presents significance (TABLE 4). However, for the SNP rs16892766 there was no evidence, given the low frequency in both cases (0.004) and controls (0.002).

**TABLE 3.**
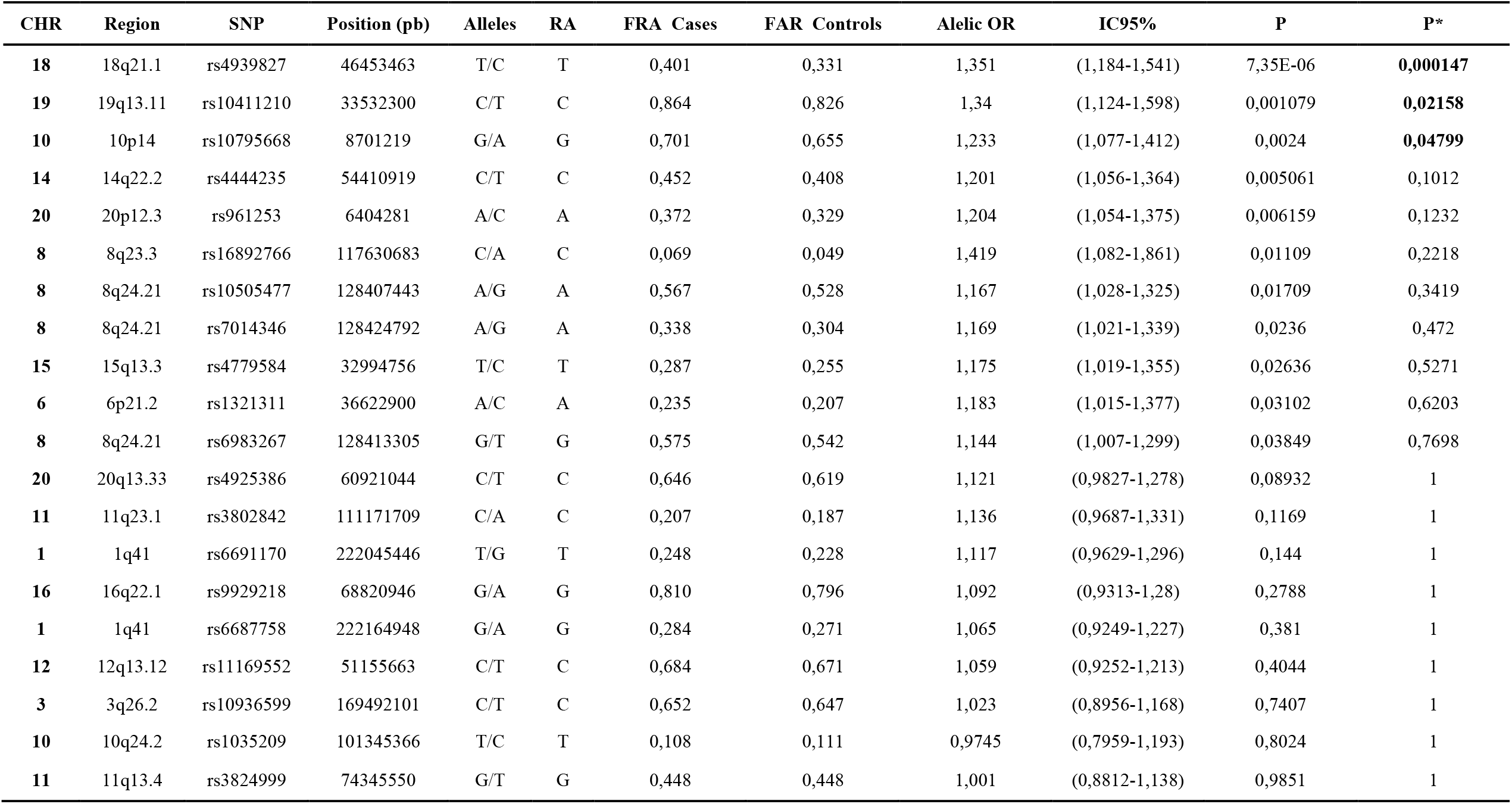
Allele frequencies and ORs in CRC cases and controls for the 20 SNPs analyzed. Allelic association test based on the genotypes of the markers, highlighting 11 nominally associated SNPs and three that passed the bonferroni test; abbreviations CHR=Chromosome, bp=base pair, RA=reported risk allele, FRA=frequency of risk allele, OR=Odds ratio, CI=Confidence interval, P=nominal p-value, obtained using a standard case association test /control (X2), P*= corrected p-value (Bonferroni).

| CHR | Region | SNP | Position (pb) | Alleles | RA | FRA Cases | FAR Controls | Alelic OR | IC95% | P | P* |
| --- | --- | --- | --- | --- | --- | --- | --- | --- | --- | --- | --- |
| 18 | 18q21.1 | rs4939827 | 46453463 | T/C | T | 0,401 | 0,331 | 1,351 | (1,184-1,541) | 7,35E-06 | **0,000147** |
| 19 | 19q13.11 | rs10411210 | 33532300 | C/T | C | 0,864 | 0,826 | 1,34 | (1,124-1,598) | 0,001079 | **0,02158** |
| 10 | 10p14 | rs10795668 | 8701219 | G/A | G | 0,701 | 0,655 | 1,233 | (1,077-1,412) | 0,0024 | **0,04799** |
| 14 | 14q22.2 | rs4444235 | 54410919 | C/T | C | 0,452 | 0,408 | 1,201 | (1,056-1,364) | 0,005061 | 0,1012 |
| 20 | 20p12.3 | rs961253 | 6404281 | A/C | A | 0,372 | 0,329 | 1,204 | (1,054-1,375) | 0,006159 | 0,1232 |
| 8 | 8q23.3 | rs16892766 | 117630683 | C/A | C | 0,069 | 0,049 | 1,419 | (1,082-1,861) | 0,01109 | 0,2218 |
| 8 | 8q24.21 | rs10505477 | 128407443 | A/G | A | 0,567 | 0,528 | 1,167 | (1,028-1,325) | 0,01709 | 0,3419 |
| 8 | 8q24.21 | rs7014346 | 128424792 | A/G | A | 0,338 | 0,304 | 1,169 | (1,021-1,339) | 0,0236 | 0,472 |
| 15 | 15q13.3 | rs4779584 | 32994756 | T/C | T | 0,287 | 0,255 | 1,175 | (1,019-1,355) | 0,02636 | 0,5271 |
| 6 | 6p21.2 | rs1321311 | 36622900 | A/C | A | 0,235 | 0,207 | 1,183 | (1,015-1,377) | 0,03102 | 0,6203 |
| 8 | 8q24.21 | rs6983267 | 128413305 | G/T | G | 0,575 | 0,542 | 1,144 | (1,007-1,299) | 0,03849 | 0,7698 |
| 20 | 20q13.33 | rs4925386 | 60921044 | C/T | C | 0,646 | 0,619 | 1,121 | (0,9827-1,278) | 0,08932 | 1 |
| 11 | 11q23.1 | rs3802842 | 111171709 | C/A | C | 0,207 | 0,187 | 1,136 | (0,9687-1,331) | 0,1169 | 1 |
| 1 | 1q41 | rs6691170 | 222045446 | T/G | T | 0,248 | 0,228 | 1,117 | (0,9629-1,296) | 0,144 | 1 |
| 16 | 16q22.1 | rs9929218 | 68820946 | G/A | G | 0,810 | 0,796 | 1,092 | (0,9313-1,28) | 0,2788 | 1 |
| 1 | 1q41 | rs6687758 | 222164948 | G/A | G | 0,284 | 0,271 | 1,065 | (0,9249-1,227) | 0,381 | 1 |
| 12 | 12q13.12 | rs11169552 | 51155663 | C/T | C | 0,684 | 0,671 | 1,059 | (0,9252-1,213) | 0,4044 | 1 |
| 3 | 3q26.2 | rs10936599 | 169492101 | C/T | C | 0,652 | 0,647 | 1,023 | (0,8956-1,168) | 0,7407 | 1 |
| 10 | 10q24.2 | rs1035209 | 101345366 | T/C | T | 0,108 | 0,111 | 0,9745 | (0,7959-1,193) | 0,8024 | 1 |
| 11 | 11q13.4 | rs3824999 | 74345550 | G/T | G | 0,448 | 0,448 | 1,001 | (0,8812-1,138) | 0,9851 | 1 |

**TABLE 4:** Dominant or recessive inheritance models in 5 of the most associated SNPs with CRC in Colombia.

| **Marcador** | rs4939827 | rs10411210 | rs10795668 | rs4444235 | rs16892766 |
| --- | --- | --- | --- | --- | --- |
| **POS** | 18(46453463) | 19(33532300) | 10(8701219) | 14(54410919) | 8(117630683) |
| **P-DOM** | 2.11E-06 | 0.1627 | 0.1066 | 0.01713 | NA |
| **P-REC** | 0.0374 | 0.001264 | 0.002373 | 0.02465 | NA |
| **OR<sub>HET</sub>**<br>**(CI95%)** | 1.529<br>(1.259-1.857) | 1.205<br>(0.658-2.206) | 1.117<br>(0.8187-1.524) | 1.201<br>(0.9804-1.472) | 1.419<br>(1.066-1.89) |
| **P<sub>HET</sub>** | 1.81E-05 | 0.5461 | 0.4849 | 0.07688 | 0.01653 |
| **OR<sub>HOM</sub>**<br>**(CI95%)** | 1.646<br>(1.245-2.175) | 1.644<br>(0.9129-2.959) | 1.445<br>(1.061-1.969) | 1.467<br>(1.123-1.918) | 2.123<br>(0.3879-11.62) |
| **P<sub>HOM</sub>** | 0.0004613 | 0.09764 | 0.01969 | 0.004965 | 0.3853 |

Significance is shown for the models tested and also the ORs for the homozygous or heterozygous status of the risk allele with the 95% CIs and P values. Abbreviations: POS= position in bp, DOM= p value in the model dominant, REC= p-value in the recessive model, OR<sub>HET</sub>= Odds ratio for heterozygote, P<sub>HET</sub>= p-value for OR of heterozygote, OR<sub>HOM</sub>= Odds ratio for homozygote, P<sub>HOM</sub>= p-value for OR of homozygote.

We also detected interactions between socioeconomic status and educational level in relation to CRC risk. In general, these two covariates presented significant effects, particularly the middle stratum compared to the low one (OR= ∼ 0.61) and the middle or higher educational level (compared to the low one), (OR= ∼1.4 and ∼2.6 respectively). Within the principal components, PC2 was significant in the regressions, which is not surprising given that it is highly correlated with European ancestry (r= -0.83, p < 2.2e-16) and also with Native American (r = 0.99, p < 2.2e-16), which does not happen with PC1, which is correlated with African ancestry (r= -0.99, p < 2.2e-16). Given the correlations between PCs and global ancestries, we only take one type into account for association with CRC risk, but not the two types of variables (based on VIF). The previous results suggest that CRC susceptibility is explained in part by the known SNPs for CRC risk, but also the ancestral component is relevant as was evidenced with PC2 association.

### Polygenic models based on the 20 SNPs

In order to evaluate the cumulative effect of risk alleles, a risk score per individual was counted (FIGURE 2). The distribution of the risk alleles in the cases and controls from Colombia depict a change around 10 risk alleles, when the cases begin to show a greater accumulation, measured as the percentage of carrier individuals presenting an OR > 1.0. (FIGURE 2b). In general, the number of risk alleles (±SD) measured in cases (9.7 ± 2.6) and controls (8.9 ± 2.5) was significantly different (t-test p=4.618e-12 or permutation =2e-04).

**FIGURE 2:**
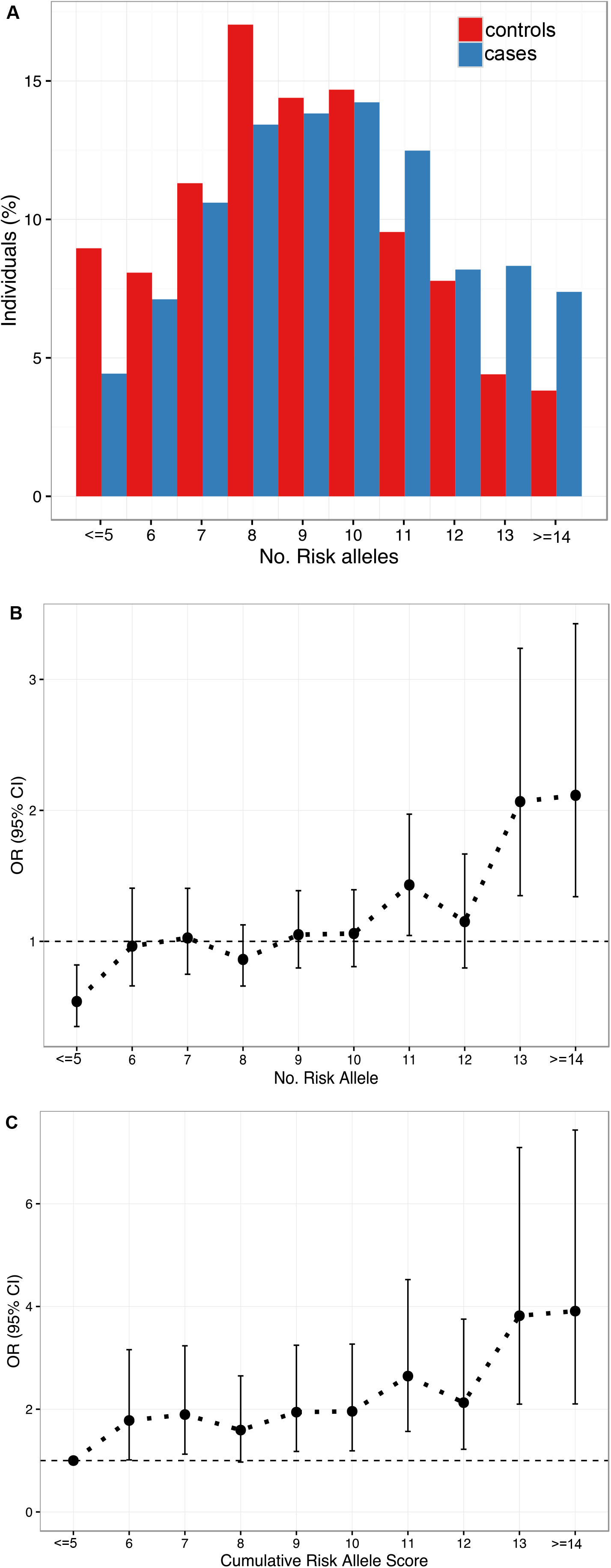
Models based on the risk allele score for the 11 nominally associated SNPs (rs4939827, rs10411210, rs10795668, rs4444235, rs961253, rs16892766, rs10505477, rs7014346, rs4779584, rs13211 and rs698). Total range of levels from 2 to 17 alleles, basal level 5 alleles or less (<=5), upper level 14 or more (>14). a) Distribution of the risk score in cases and controls; b) Odds ratios (95% CI) calculated for each category, c) cumulative effect adding one risk allele at the same time/category, compared with the baseline or reference level (5 or less =1).

The risk score ranged from 2 to 17 risk alleles, establishing the lower and upper limits at 5 and 14 alleles respectively, in order to obtain reliable estimates in the tails of the distribution. The increased risk due to the gradual accumulation of susceptibility alleles in a model where each successive level is compared with the baseline (FIGURE 2c, TABLE 5) indicates that there is a 5.39-fold increased risk when an individual carries 14 or more risk alleles, compared to that of 5 alleles.

**TABLE 5:** Cumulative effect of the number of risk alleles on the susceptibility to CRC.

| **risk score** | **Cases (n)** | **Controls (n)** | **% Cases** | **% Controls** | **OR** | **IC (95% IC)** | **P** |
| --- | --- | --- | --- | --- | --- | --- | --- |
| **<=5** | 86 | 35 | 3.89 | 9.21 | 1 (Reference) |  |  |
| **6** | 75 | 63 | 7.01 | 8.03 | 2.064 | (1.237-3.482) | 0.005949 |
| **7** | 108 | 95 | 10.57 | 11.56 | 2.161 | (1.346-3.521) | 0.00165 |
| **8** | 161 | 116 | 12.9 | 17.24 | 1.77 | (1.125-2.828) | 0.014898 |
| **9** | 135 | 124 | 13.79 | 14.45 | 2.257 | (1.431-3.615) | 0.000561 |
| **10** | 138 | 128 | 14.24 | 14.78 | 2.279 | (1.448-3.645) | 0.000458 |
| **11** | 88 | 116 | 12.9 | 9.42 | 3.239 | (2.017-5.285) | 1.66E-06 |
| **12** | 72 | 74 | 8.23 | 7.71 | 2.525 | (1.526-4.235) | 0.000367 |
| **13** | 40 | 80 | 8.9 | 4.28 | 4.914 | (2.87-8.575) | 1.12E-08 |
| **>=14** | 31 | 68 | 7.56 | 3.32 | 5.39 | (3.052-9.731) | 1.16E-08 |

The ORs (95% CI) and p values were obtained using a logistic regression model (dummy contrast) in which the reference level is the category of 5 or fewer risk alleles, with which the other levels were compared.

### Relationships between low penetrance CRC susceptibility loci and some clinicopathological characteristics

Previous works by the research group have analyzed the clinical and pathological information in the CHIBCHA samples for Colombia, in order to detect associations between them (Bohórquez et al., 2016). In the present work we addressed the relationship between CRC susceptibility SNPs with some clinicopathological variables through logistic regression analysis. The cases were divided into subgroups according to family history, defined as cases with at least one first-degree relative with CRC -FH-(positive or negative, n=747), age of onset stratified into two groups -EIE-(< 50, or >50, n=747), or as a continuous variable -EIC-, degree of tumor differentiation -TG-(high or low, n=614), tumor location -TL-(proximal colon, distal colon and rectum, n=332). Additionally, covariates were included: gender, socioeconomic status -SE-, education -ED- and European ancestry -EUR- or -NAM-. Two SNPs were found to be associated with family history: rs10795668 (10p14) (p =0.0306, OR= 3.3 CI: 1.246-14.5) and rs1035209 (10q24.2) (p=0.0464, OR=0.548, CI: 0.29-0.96); likewise, the high degree of tumor dedifferentiation was found to be associated with an allele in the SNP rs1321311 (6p21.2) (p=0.019, OR=2.18, CI: 1.14-4.17), and the onset of CRC before the age of 50 was negatively associated with rs7014346 (OR=0.42, CI: 0.27-0.65, p= 0.000097). Regarding to the tumor location, having three categories, paired regressions were performed and the rs7014346 was associated with right-sided CRC (p= 0.026, OR= 2.083 CI: 1.108-3.98), while rs132131 was associated with the Left-sided CRC (p= 0.041, OR=3.23 CI: 1.1-10.6). Considering an interaction analysis between SNPs, a significant value was found between rs7014346 and rs1321311 for tumors located on the right side (p= 0.0131).

## DISCUSSION

Several genetic aspects were analyzed in a sample of the Colombian population in relation to CRC risk. A sample set of 955 cases and 968 controls was recruited for the CHIBCHA consortium in Colombia; from which global genomic data of about one million SNPs were obtained. In general, the CHIBCHA sample came mostly from populated centers located in the Andean Region, were approximately 30 million inhabitants reside, nearly∼70.1% of the total Colombian population (DANE, 2018).

The Colombian Andes are a geographical extension with climatic and landscape variations that have influenced the migration and distribution of human groups throughout its territory, with cultural differences that in turn, may be linked to demographic history such as the initial native American populations originally stablished in pre-Columbian times, and the characteristics of the Spanish immigrants who arrived in colonial times. Therefore, the genetic characteristics of the modern populations in Colombia are a consequence of initial conditions such as the diversity and density of the indigenous populations, their persistence over time and the patterns of admixture that occurred between native groups, the Europeans and Africans brought to Colombia from the respective Spanish colonies in Africa. This is particularly interesting given the variations in the density of ethnic groups, both indigenous and Afro-Colombian, between the different departments in Colombian Andes, which can be related to variations in the genetic ancestral background (DANE, 2018).

The genetic variation observed in the Colombian Andean populations influenced by the aforementioned factors, can be extended to disease susceptibility. For example, CRC has a high incidence in one of the parental populations -European Iberian-, but lower in Colombian population which can be due to the lost/conservation dynamic of CRC risk-related variation during admixture process as a consequence of stochastic processes (drift) during colonial times. On the other hand, the contribution to CRC from Native Americans is little known (or absent). Evidence of this process can be found in the epidemiological behavior of the disease, for example, some studies carried out by the Center for Disease Control and Prevention in the United States -CDC-, on the incidence of CRC in some indigenous populations, have estimated an incidence between 58 and 34/100,000 (between the years 1999-2013), lower than that observed for populations of Caucasian origin in the same study (68-44/100,000), while in populations of Hispanic origin, the incidence was intermediate (58-50/100,000) (Perdue et al., 2014; White et al., 2014).

In general, when two parental continental populations present different incidence rates for a complex disease and they intercross, intermediate disease rates can be present in admixed people (Risch & Merikangas, 1996); for example, in CRC the highest incidence is found in Caucasian populations and the lowest in Native Americans, but intermediate in Latin American mestizo populations. As a consequence of admixture, mosaic chromosomes arise because of the meiotic recombination process along the generations, with segments that can be traced to the different contributing parental populations, and this originates new loci relationships.

### Population genetic structure, ancestry and CRC risk

Cases and controls had a predominantly Native American and European genetic contribution and low differentiation between them. Understanding the genetic structure in human populations is from fundamental interest for biomedical, forensic, and anthropological studies. For example, in genetic association studies, variation in the case-control sample may influence the associations between previously reported markers of susceptibility and disease (Choudhry et al., 2006; Price et al., 2010). Perhaps the low level of genetic Fst between cases and controls, we included PCs or ancestry admixture proportions in regression models in order to correct for bias due to the detected level of subpopulation genetic structure.

In the present study a higher proportion of the European ancestral component was observed in the cases compared to the controls and *vice versa* for the Native American proportion; both ancestries being highly significant in the logistic regression models. These findings support the inclusion of the ancestral component as explanatory variable for the CRC prediction in mixed populations such as the Colombian population; we particularly observed a positive association for European population (risk factor) and a negative one for the Native American (protective?). The European admixture showed levels of population strata with differences in risk between the levels of European ancestry analyzed, for example an European ancestry > 50% is associated with the risk for CRC. These observations suggest that one or more alleles highly frequent in the European population and present in the Colombian Andean region, could increase the risk for CRC, although this association can also be due to environmental factors that were not considered, which are common in the region. Gene-environment interactions may be also explaining the differences, for instance an environmental factor that increases the CRC risk only in the presence of a particular genotype which could be in high frequency within the European population. Therefore, only the identification of particular loci that contribute to the difference between Europeans and Native Americans in relation to CRC risk can confirm that the observed difference between cases and controls is genetic and not environmental.

The presumptive protective effect of Native American ancestry for CRC risk suggests that these ethnic groups may be less likely prone to develop the disease; however, studies and/or reports on the incidence in indigenous populations are scarce. Although there are no updated registries for indigenous populations in Latin America, recent reviews pinpoint the most frequent types of cancer include cervical, stomach, prostate, and gallbladder cancers (Moore, Forman, Piñeros, Fernández, de Oliveira Santos, et al., 2014). In previous studies on the incidence of CRC, a lower incidence has been reported in Native Alaskan communities between the years 1999-2013 (Center for Disease Control and Prevention, 2016; Perdue et al., 2014), and in general in native populations. Low CRC incidence and mortality rates have been observed in North America, which may be partly due to the type of diet, for example with a higher amount of fiber (Cobb & Paisano, 1998).

In Colombia, reports from *Ministerio de Seguridad Social* about the health profile in Colombian indigenous population in 2016 (Sandoval-Castaño, 2016), reported neoplasms as the fourth cause of death (13.47% of all deaths), gastric cancer being the most represented. Specific data for CRC is not mentioned, but a few studies in populations from the Amazon and Cauca, indicate a high risk for cervical cancer among Amazon women and gastric cancer is high in Cauca men; in both studies, the density of the indigenous population is among the highest in the country (Moore, Forman, Piñeros, Fernández, Oliveira Santos, et al., 2014).

Analysis in 20 known CRC risk SNPs resulted in 11 associated markers located in the regions: 18q21.1, 19q13.11, 10p14, 14q.2.2, 20p12.3, 8q23.3, 6p21.2, 15q13.3 and 8q24.21, some SNPs presented greater association than others. Although each marker individually contributed a low risk, in polygenic models they presented a strong cumulative effect for CRC risk, which is important at the population level because it helps to identify risk groups within the cases, including family history (Malcolm G. Dunlop et al., 2012; Thomas et al., 2020; Weigl et al., 2018). Individuals with more than 10 risk alleles could eventually be candidates for screening tests; however, it is necessary to assess functional role of the association of these variants in future studies.

When adjusting the association models including socioeconomic level, department of birth, education, gender and PCs, only five markers remained significant, in the regions: 18q21.1 (rs4939827), 19q13.11 (rs10411210), 10p14 (rs10905436), 14q.2.2 () and 8q23.3 (). Some of these chromosomal regions harbor genes involved in key steps from the cell metabolism or regulatory sequences: SMAD7 (Mothers against decapentaplegic homolog 7) (18q21.1) (Broderick et al., 2007; Zeng et al., 2016); Rho GTPasa (RHPN2) (19q13.11) (Bellovin et al., 2006; Chang et al., 2006; Peck et al., 2002); transcriptional regulatory sequences at 8q23.3, encompasing the predicted transcripts: EIF3H and C8orf53 (8q23.3) (Okamoto et al., 2003; Savinainen et al., 2004).

Taken together, these data provide support for the consolidation of common genetic causes of CRC susceptibility, when comparing different populations such as the Colombian and the European. Comparatively, the degree of risk observed in Colombia for the associated SNPs in all cases followed the same trend observed in Caucasian populations, only rs4939827 presented a higher OR. The confidence intervals were wider in the present study given the sample size. The 5 most associated markers followed genetic patterns of recessiveness and therefore the highest risk was detected in the homozygous state.

## Data Availability

All data produced in the present work are contained in the manuscript

## CONCLUSIONS

The cases and controls were heterogeneous given their genetic admixture. Variations at the individual level in the Native American, European, and African ancestry proportions were observed, however the substructure was very low and did not affect the genetic associations observed when this variable was included or not into the statistical models. European ancestry may be involved in the CRC risk in Colombian population, given the history of Iberian admixture since pre-Columbian times.

Eleven out of 20 CRC risk markers were nominally replicated in the Colombian population, located in 9 chromosomal regions: 8q23.3 (rs16892766), 8q24.21 (rs6983267, rs10505477, rs7014346), 10p14 (rs10795668), 14q22.2 (rs4444235), 15q13.3 (rs4779584), 18q21.1 (rs4939827), 20p12.3 (rs961253); 19q13.11 (rs10411210) and 6p21.2 (rs1321311); presenting Odd ratios between 1.14 and 1.35. Five of them remained significant after adjusting the models for the population structure and socioeconomic characteristics of the sample: 18q21.1, 19q13.11, 10p14, 14q22.2, 8q23.3.

Two socioeconomic characteristics such as educational level and social class did affect the crude associations observed, reducing the number of significant markers from 11 to 5; Therefore, part of the differences observed between cases and controls for 6 of the markers could initially be due to the stratification of the sample due to the characteristics of the sampling process.

CHIBCHA (study of hereditary cancer in Europe and Latin America) collaborators include: Ma. Magdalena Echeverry de Polanco, Mabel Elena Bohórquez, Rodrigo Prieto, Angel Criollo, Carolina Ramírez, Ana Patricia Estrada, Jhon Jairo Suárez (Grupo de Citogenética Filogenia y Evolución de Poblaciones, Universidad del Tolima, Colombia); Augusto Rojas Martinez (Center for Research and Development in Health Sciences, Universidad Autónoma de Nuevo León, Monterrey, Mexico); Silvia Rogatto, Samuel Aguiar Jnr, Ericka Maria Monteiro Santos (Department of Urology, School of Medicine, UNESP - São Paulo State University, Botucatu, Brazil); Monica Sans, Valentina Colistro, Pedro C. Hidalgo, Patricia Mut (Department of Biological Anthropology, College of Humanities and Educational Sciences, University of the Republic, Magallanes, Montevideo, Uruguay); Angel Carracedo, Clara Ruiz Ponte, Ines Quntela Garcia (Fundacion Publica Galega de Medicina Xenomica, CIBERER, Genomic Medicine Group-University of Santiago de Compostela, Hospital Clinico, Santiago de Compostela, Galicia, Spain); Sergi Castellvi-Bel (Department of Gastroenterology, Institut de Malalties Digestives i Metabòliques, Hospital Clínic, Centro de Investigación Biomédica en Red de Enfermedades Hepáticas y Digestivas, IDIBAPS, University of Barcelona, Barcelona, Catalonia, Spain); Manuel Teixeira (Department of Genetics, Portuguese Oncology Institute, Rua Dr, António Bernardino de Almeida, Porto, Portugal).

