## Supplementary material for "Colorectal Cancer Risk and Ancestry in Colombian admixed Populations"

Supplementary figure 1: Ancestral proportions distribution goodness of fit in colorectal cancer cases and controls by using 87K SNP dataset.

Cullen and Frey plots are shown with the kurtosis and skewness coefficients, adjusting the known distributions. It is observed admixture proportions (blue dot) and the bootstraped values (yellow). Native (a) and European (b) ancestry tend to fit the logistic distribution, very close to normal, while African (c) is observed with a different pattern

(gamma). Below each Cullen and Frey plot, the observed densities (black line) and the fitted theoretical distribution (red line) are shown.

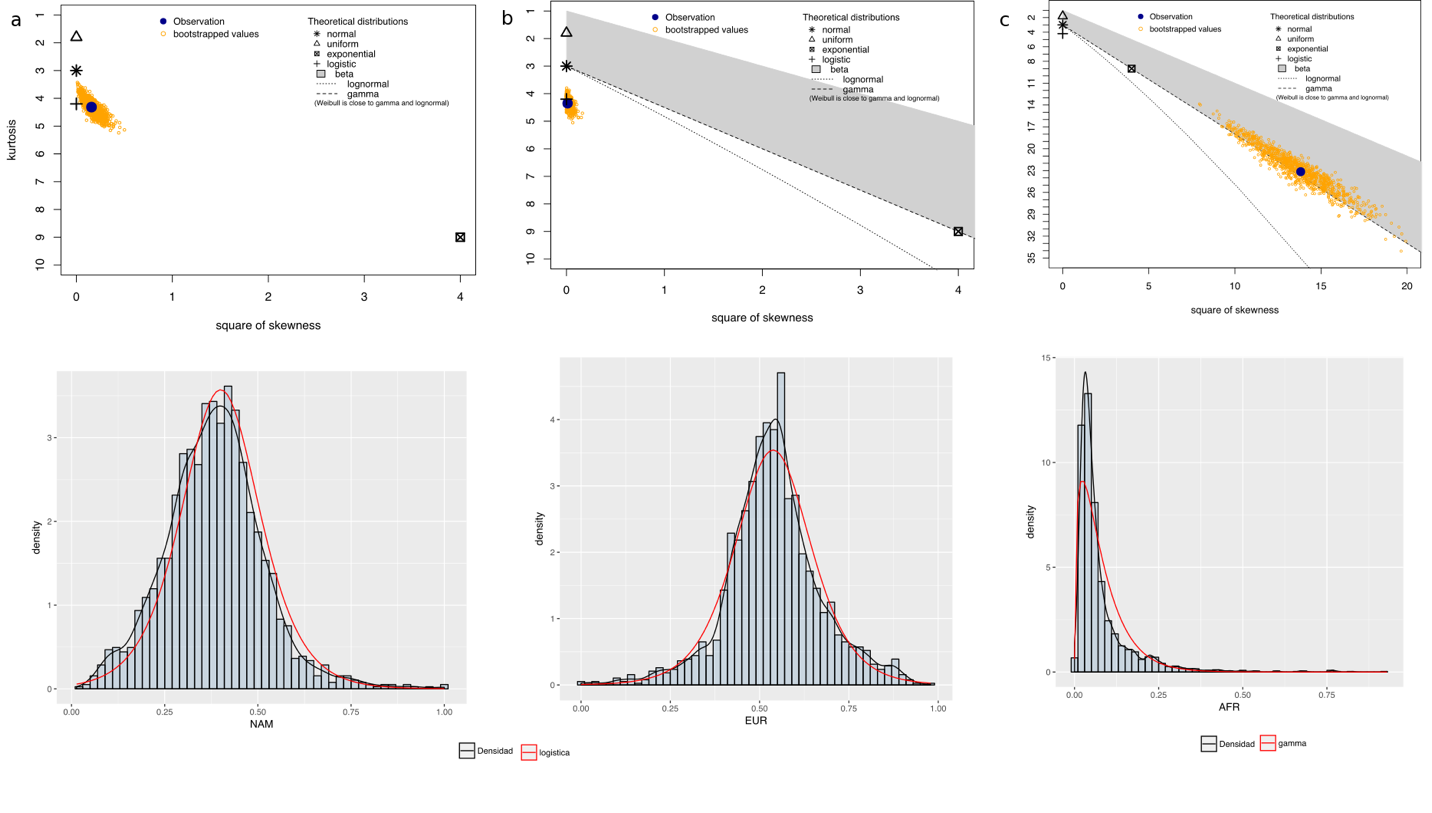

Supplementary Table 1: Logistic regressions for five nested models to predict CRC risk (cases/controls) in terms of ancestry. The phenotype was assumed as the dependent variable, and the native ancestry as the main regressor variable. The univariate model is shown in increasing order, that is, only using European ancestry (EUR) as the regressor variable. Each model with the addition of one covariate at a time (gender -GE-, department -DEP-, socioeconomic stratum - EST-, education -EDU-). The table reports the logarithm of the likelihood of each model (LogLik), the estimated value of the coefficient (beta), the standard error, the value of the z statistic, and p-val.

| **Model** | **Variable** | **Coeficient** | **Estandar error** | **Z** | **Pr(>\|z\|)** |
| --- | --- | --- | --- | --- | --- |
| **m1. status ~ EUR** | (Intercept) | -0,6893018 | 0,2235106 | -3,083,978 | 0,002042525 |
| LogLik: -1030,9 | **EUR** | 14,526,075 | **0,3984763** | 3,645,405 | **0,000266971** |
| **m2. status ~ EUR + GE** | (Intercept) | -0,5739953 | 0,2293603 | -2,502,592 | 0,01232875 |
| LogLik:-1028,4 | **EUR** | 14,873,788 | 0,39934 | 3,724,592 | 0,000195631 |
|  | GE(women) | -0,2356554 | 0,1052472 | -2,239,067 | 0,025151589 |
| **m3. status ~ EUR + GE + DEP** | (Intercept) | -0,916488 | 0,2874985 | -318,780,081 | 0,001433593 |
| LogLik:-1021,4 | **EUR** | **185,985,268** | **0,4415393** | 421,220,223 | **2.53E+00** |
|  | GE(women) | -0,22379373 | 0,106329 | -210,472,992 | 0,035314824 |
|  | Boyacá | -0,02233241 | 0,2413033 | -0,09254912 | 0,926261763 |
|  | Costa | 0,25713675 | 0,3715902 | 0,6919901 | 0,488943544 |
|  | Cundinamarca | 0,03022555 | 0,180116 | 0,16781163 | 0,866731473 |
|  | Eje cafetero | -0,11639934 | 0,2473945 | -0,47050101 | 0,637997111 |
|  | Huila | -0,08210052 | 0,2227498 | -0,36857732 | 0,712442803 |
|  | Nariño | 0,37811821 | 0,23849 | 15,854,681 | 0,11286002 |
|  | Santanderes | 0,34634656 | 0,2741163 | 126,350,226 | 0,206408735 |
|  | Tolima | 0,42756921 | 0,1508163 | 283,503,231 | 0,004582105 |
|  | Valle y Cauca | 0,35155687 | 0,3588164 | 0,97976818 | 0,327200561 |
| **m4. status ~ EUR + GE + DEP + EST** | (Intercept) | -0,73642801 | 0,293229 | -251,144,366 | 0,012023848 |
| LogLik:-1015,6 | **EUR** | **173,939,282** | **0,4600407** | 378,095,451 | **0,000156228** |
|  | GE(women) | -0,22551666 | 0,1067386 | -211,279,287 | 0,0346185 |
|  | Boyacá | -0,07502561 | 0,2441089 | -0,30734483 | 0,758580917 |
|  | Costa | 0,13961217 | 0,3785018 | 0,36885472 | 0,712236015 |
|  | Cundinamarca | 0,01577278 | 0,181257 | 0,08701887 | 0,930656511 |
|  | Eje cafetero | -0,13545661 | 0,2492506 | -0,54345557 | 0,586816176 |
|  | Huila | -0,17210948 | 0,2293632 | -0,75037982 | 0,453025984 |
|  | Nariño | 0,31621034 | 0,2396873 | 131,926,216 | 0,187081483 |
|  | Santanderes | 0,3333597 | 0,2752127 | 121,128,043 | 0,225787947 |
|  | Tolima | 0,4080278 | 0,1529717 | 266,734,235 | 0,007645375 |
|  | Valle y Cauca | 0,3176599 | 0,3612195 | 0,87940951 | 0,379179278 |
|  | EST-middle | -0,26040313 | 0,1141608 | -228,102,127 | 0,022547188 |
|  | EST-high | 0,45272667 | 0,2594072 | 174,523,583 | 0,080943827 |
| **m5. status ~ EUR + GE + DEP + EST + EDU** | (Intercept) | -0,64006873 | 0,2959652 | -216,264,865 | 0,03056821 |
| LogLik:-1003.1 | **EUR** | 144,539,835 | **0,4682979** | 308,649,304 | **0,002025327** |
|  | GE(women) | -0,15855907 | 0,1084598 | -14,619,161 | 0,1437642 |
|  | Boyacá | -0,14893449 | 0,2461339 | -0,60509543 | 0,5451156 |
|  | Costa | -0,15089474 | 0,3870932 | -0,38981501 | 0,6966733 |
|  | Cundinamarca | -0,1046917 | 0,1849877 | -0,56593858 | 0,5714355 |
|  | Eje cafetero | -0,18718443 | 0,2507645 | -0,74645502 | 0,4553926 |
|  | Huila | -0,27764032 | 0,2317396 | -119,807,008 | 0,2308897 |
|  | Nariño | 0,17498077 | 0,2434326 | 0,71880579 | 0,4722606 |
|  | Santanderes | 0,22891639 | 0,2785808 | 0,82172355 | 0,4112343 |
|  | Tolima | 0,26014372 | 0,1568909 | 165,811,865 | 0,09729352 |
|  | Valle y Cauca | 0,2257528 | 0,3643078 | 0,61967607 | 0,5354711 |
|  | EST-middle | -0,46057403 | 0,1223729 | -376,369,291 | 0,0001674225 |
|  | EST-high | 0,01756536 | 0,2766457 | 0,06349408 | 0,9493731 |
|  | EDU-middle | 0,36341426 | 0,1255301 | 289,503,635 | 0,003791146 |
|  | EDU-high | 0,90141173 | 0,1905402 | 473,082,175 | 0,000002236128 |
| Status: case/control | |  |  |  |  |

Complete List CHIBCHA consortium:

CHIBCHA (study of hereditary cancer in Europe and Latin America) collaborators

include: Ma. Magdalena Echeverry de Polanco, Mabel Elena Bohórquez, Rodrigo Prieto,

Angel Criollo, Carolina Ramírez, Ana Patricia Estrada, Jhon Jairo Suárez (Grupo de

Citogenética Filogenia y Evolución de Poblaciones, Universidad del Tolima, Colombia);

Augusto Rojas Martinez (Center for Research and Development in Health Sciences,

Universidad Autónoma de Nuevo León, Monterrey, Mexico); Silvia Rogatto, Samuel Aguiar

Jnr, Ericka Maria Monteiro Santos (Department of Urology, School of Medicine, UNESP -

São Paulo State University, Botucatu, Brazil); Monica Sans, Valentina Colistro, Pedro C.

Hidalgo, Patricia Mut (Department of Biological Anthropology, College of Humanities and

Educational Sciences, University of the Republic, Magallanes, Montevideo, Uruguay); Angel

Carracedo, Clara Ruiz Ponte, Ines Quntela Garcia (Fundacion Publica Galega de Medicina

Xenomica, CIBERER, Genomic Medicine Group-University of Santiago de Compostela,

Hospital Clinico, Santiago de Compostela, Galicia, Spain); Sergi Castellvi-Bel (Department

of Gastroenterology, Institut de Malalties Digestives i Metabòliques, Hospital Clínic, Centro

de Investigación Biomédica en Red de Enfermedades Hepáticas y Digestivas, IDIBAPS,

University of Barcelona, Barcelona, Catalonia, Spain); Manuel Teixeira (Department of

Genetics, Portuguese Oncology Institute, Rua Dr, António Bernardino de Almeida, Porto,

Portugal).
